# EarlyDetect: Development of tool for early detection of stomach adenocarcinoma based on blood-lipid profile as clinicopathological feature

**DOI:** 10.1101/2023.02.14.23285899

**Authors:** Om Prakash, Feroz Khan

## Abstract

**Background:** Clinicopathological features are used for detection of diseases. Early detection of cancer can be significance for understanding the behavior of disease.

**Results:** We developed a tool to observe stomach adenocarcinoma in reference of blood-lipid profile. Background of the tool is based on the study made on RNAseq expression analysis of stomach adenocarcinoma. Raw data for study was collected as gene-expression profile from population of cancer-vs-normal. A series of studies performed including: differential gene expression analysis, plasma proteome mapping, extraction of gene-signature enriched with LSTM system model, AI-guided simulation of systems model of gene network, and AI-guided mapping with blood lipid profile to develop R-Shiny web-application.

**Conclusion:** ‘EarlyDetect’ is freely available at https://csir-icmr.shinyapps.io/EarlyDetect/. The tool can be utilized for (i) virtual observation of impact of different combinations of lipid profile in cancer progression; (ii) early detection of cancer state for new comer patients.

## INTRODUCTION

Blood lipid is a clinicopathological feature which is used to observe disease status. Lipid profile is also useful in detection of cancer as in case of: colorectal, breast, lung, ovarian and prostate cancers etc [1]. Lipid analogs are also known as key regulators of tumorigenesis [2]. Such studies motivate for observation of cancer in relation of lipid profile [3]. Lipid profile shows mutual influence due to drug response. As, hormonal treatment of breast cancer reduces the oestrogen level, which further reduces the blood lipid level [4]. Therefore, blood profile along with percentage of dyslipidemia are observe before chemotherapy [5]. Research is being performed on association between blood lipid level and cancer specific mortalities. Such studies work on variation in blood lipid profile including ‘total cholesterol’, triglycerides, and HDL/LDL level [6]. Blood lipid level is observed along with blood glucose and extent of inflammation, as in case of ovarian cancer [7]. All these studies suggest that abnormal blood lipid profiles contain association with occurrence of cancer [8]. Therefore, blood lipid profile is regularly estimated, if cancer patient is under endocrine therapies [9]. Other studies like phospholipid level & peroxidation estimation are performed to understand the malignant neoplasma as in case of breast & uterine cancer [10]. Drug response studies on blood profile are also available for tamoxifen, endoxifen and 4-hydroxyTamoxifen [11]. In totality, potential correlation exists between blood lipid and cancer [12]. Diagnostic values are also considerable in combination of tumor markers and blood lipid [13]. These studies support and prepare a ground for establishing relationship between genotype (tumor markers) and phenotype (blood lipid profile) for observation of import of variation in blood lipid and the cancer status.

RNA expressions are used in several areas of clinical diagnosis [14], including cancer [15]. Categories of RNA as: miRNA [16], circRNA expression [14], long noncoding RNA expression [15], mRNA expression [17] are used in diagnosis. Similar to DNA & protein biomarkers, RNA biomarkers are also potential to discriminate diseased-vs-normal conditions [18]. Therefore, transcriptome sequencing are used in diagnosis of disorders [19]. Transcriptome RNA expressions are used for identification of targetable disease regulatory networks for disease diagnosis [20]. RNA based studies are known with hepatocellular carcinoma [21], colorectal cancer [22], pathway detection [23], prostate tumor [24], and breast cancer [16] etc. RNA expression profile/signature are also used for early diagnosis [25, 26]. Early diagnosis is directly linked with development of biomarker signature (i.e., gene set) with population discrimination capacity [27]. Such studies provide clinically relevant results for diagnosis [28]. These studies provide a ground for accessing RNA expressions as genotypic features for early detection of cancer. Genotype-to-phenotype relation is used also for linking lipid metabolism with cancer related genes [29].

Stomach adenocarcinoma (STAD) is one of the leading reasons of deaths in the world. Lack of early detection is also major barrier behind this situation. Biomarker genes linked with extracellular matrix and platelet-derived growth factor are suggested in early detection of STAD [30]. Besides this, CDCA7-regulated inflammatory mechanism [31], angiogenesis-related lncRNAs [32], PLXNC1 an immune-related gene [33], and Ferroptosis-related gene [34] etc. are also claimed for early detection of STAD. Different stages of STAD are available through prognostic model of TCGA database [35]. Various combination of gene-signature genes is still remained to explore, which creates opportunity to reveal aspect for early detection of STAD.

In present study, blood lipid profile (as phenotype) was linked with gene signature (as genotype) derived for stomach adenocarcinoma, to predict the relative changes/ status of cancer. This process was packaged in the form of R-Shiny GUI web-application named ‘EarlyDetect’.

## MATERIALS AND METHOD

‘EarlyDetect’ establishes relationship between Phenotype and Genotype defined for disease features. It establishes AI-guided relation between blood lipid-profile (clinicopathological phenotype) and expression of gene signature. ‘EarlyDetect’ works as tool for making decision for any unknown sample with blood lipid-profile. Workflow for tool is described as:

### Phenotype for EarlyDetect

Although cancer is seeded at molecular level, but it can be featured by clinicopathological phenotypes. Cancer-start-up alters gene expressions. Identification of differentially expressed genes is always a relevant question. Present study deals with establishment of relationship between set of gene expressions (gene signature) and clinicopathological phenotypic character for designing of tool. Prior studies indicate towards the influence of blood lipid-profile (phenotype) during the start-up of cancer. Considering prior studies, blood-lipid profiles featured as phenotype for the study. General blood lipid profile used in this study, is: Total Cholestrol: 200-239(mg/dL); LDL Cholestrol: 130-159 (mg/dL); HDL Cholestrol: 40-60 (mg/dL); Triglycerides: 150-199 (mg/dL); Non-HDL-C: 130-159 (mg/dL); and TG to HDL ratio: 3.0-3.8 (mg/dL).

### Genotype for EarlyDetect

TCGA database provides normalized RNAseq expression for multiple cancer types. TCGA contains 408 independent tumor samples and 211 matching normal tissue samples for Stomach adenocarcinoma (STAD) showing more than 29K genes. Log-fold change in gene expression, above significant threshold, filter outs differentially expressed genes (DEGs). Volcano plot represents DEGs graphically. Since DEGs also display into blood samples; therefore, to filter DEGs, plasma proteome mapped with it. Here mapped DEGs (pDEGs) are DEGs available in blood plasma proteome. Co-expression network analysis processed pDEGs for identification of gene-signature. Details of procedure can be accessed from doi.org/10.1038/s41598-021-87037-w [36]. Gene-wise similarity correlation matrix (scor) & dis-similarity correlation matrix (dcor) presented co-expression of pDEGs. Calculation of scor & dcor takes input of normalized gene expression of pair of genes (G_i_ & G_j_). Here, ‘scor’ represents the positive correlation matrix between the genes since the larger value indicates the stronger positive correlation between the pair of genes, while ‘dcor’ represents the negative correlation between a pair of genes since the larger value indicates the stronger negative correlation between genes, which is also called the distance of a pair of genes. Network analysis used both dissimilarity and similarity matrices with values ranging between 0 & 1. Gene network contained 39 genes. pDEGs in any of these two networks can be considered as co-expressed genes; and therefore, assumed to be involved in existence of system model representing state of disease. Here similarity matrix used for further analysis.

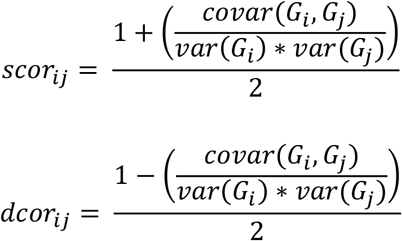

System model, based on pDEGs based on similarity matrix, performed further enrichment of co-expression of gene-set. Long-Short-Term-Memory Recurrent Neural Network (LSTM-RNN) model (implemented in R) simulated the system model. Jacobian matrix performed evaluation of stability of system model. LSTM enriched gene signatures were genotypes. Tool development utilized gene-signature.

### Evaluation of signature for capacity of population discrimination

Survival plot, between diseased & normal patients, presents discrimination capacity of gene-signature in the population. Therefore gene-signatures processed for finding survival plot, observed through Logrank p-value < 0.05 as threshold for signaificant expression.

### Linking gene signature (genotype) with blood lipid profile (phenotype)

To link genotype with phenotype, LSTM enriched gene signature mapped with lipid-profile of blood. Total 740 (370 (normal) + 370 (cancer)) data-points ranged for gene expression, mapped with lipid-profile. Since, cancer-vs-normal gene expressions are pre-classified, therefore classified gene expression for gene signature made classified mapped lipid-profile dataset. Further, mapped model developed based on the classified genotypic & phenotypic dataset sets, and further utilized for development of map-model for transformation of blood profile into gene expression. Artificial Neural Network (feed-forward backpropagation) mapped the genotype-vs-phenotype profiles. Phenotype contained 06 nodes of blood lipid-profile, while genotype contained 5 nodes of enriched pDEGs.

### Evaluation of mapped lipid profile for the capacity of population discrimination

Since genotype used to map phenotype, therefore lipid profile should also contain population discrimination capacity. Therefore, population discrimination capacity was also evaluated for mapped lipid profile. Total 740 (370 (normal) + 370 (cancer)) data-points ranged for lipid-profile mapped in reference of gene-signature.

Two class perceptron classification method utilized mapped lipid-data points for evaluating population discrimination capacity. Area-Under-Curve (AUC) of ROC plot evaluated the classification model.

### Deployment of Shiny tool ‘EarlyDetect’

R-shiny platform used to develop a web-application, which receive 06 inputs of blood lipid profile, and transform phenotype into gene expression profile with 05 outputs; and further provides output in the form of probability of cancer. The developed application deployed at Shiny-Server.

## RESULTS AND DISCUSSION

### DEGs for stomach adenocarcinoma in plasma

Total 62 differentially expressed genes found after processing 4644 genes under the thresholds as ‘Fold change (log_2_)’ of 1.5 and ‘Significance (-log_10_)’ boundary at 0.05 (Figure 2). Furthermore, plasma proteome database (http://www.plasmaproteomedatabase.org) mapping filtered 39 genes out from 62 DEGs (Table 1). These 39 genes (gene symbol marked with ‘node ID’ from 1 to 39 in table-1) processed strategically for development of gene-signature.

**Table 1.** Plasma-expressed DEGs. Differentially expressed genes mapped along plasma proteome. 39 genes, out of 62, found to be expressed in blood plasma. Considering the feasibility of patient sampling through blood, plasma proteome expressions used for identification of gene-signature. Here in table each gene symbol is marked with ‘node ID’ from 1 to 39.

| Node ID -vs- Gene | PPD ID | Gene symbol | Gene name | Entrez gene ID |
| --- | --- | --- | --- | --- |
| 1 | HPRD_03272 | AFF3 | AF4/FMR2 family, member 3 | 3899 |
| 2 | HPRD_04087 | APBB1 | Amyloid beta (A4) precursor protein-binding, family | 322 |
|  |  |  | B, member 1 (Fe65) |  |
| 3 | HPRD_00134 | APOD | Apolipoprotein D | 347 |
| 4 | HPRD_00405 | C5 | Complement component 5 | 727 |
| 5 | HPRD_03148 | CHAF1A | Chromatin assembly factor 1, subunit A (p150) | 10036 |
| 6 | HPRD_04592 | CHRD | Chordin | 8646 |
| 7 | HPRD_02363 | COL4A5 | Collagen, type IV, alpha 5 | 1287 |
| 8 | HPRD_06012 | DPP3 | Dipeptidyl-peptidase 3 | 10072 |
| 9 | HPRD_00510 | DPT | Dermatopontin | 1805 |
| 10 | HPRD_04265 | EEF1A2 | Eukaryotic translation elongation factor 1 alpha 2 | 1917 |
| 11 | HPRD_03225 | EFNA3 | Ephrin-A3 | 1944 |
| 12 | HPRD_03957 | PDK4 | Pyruvate dehydrogenase kinase, isozyme 4 | 5166 |
| 13 | HPRD_06685 | PRSS3 | Protease, serine, 3 | 5646 |
| 14 | HPRD_00340 | VCAN | Versican | 1462 |
| 15 | HPRD_03642 | NRP1 | Neuropilin 1 | 8829 |
| 16 | HPRD_06714 | SEC23A | Sec23 homolog A (S. Cerevisiae) | 10484 |
| 17 | HPRD_09843 | SLC52A3 | Solute carrier family 52, riboflavin transporter, member 3 | 113278 |
| 18 | HPRD_01418 | SERPINE1 | Serpin peptidase inhibitor, clade E (nexin, plasminogen activator inhibitor type 1), member 1 | 5054 |
| 19 | HPRD_13296 | FAM20A | Family with sequence similarity 20, member A | 54757 |
| 20 | HPRD_11762 | ZBTB16 | Zinc finger and BTB domain containing 16 | 7704 |
| 21 | HPRD_01475 | PTPN6 | Protein tyrosine phosphatase, non-receptor type 6 | 5777 |
| 22 | HPRD_01087 | LOX | Lysyl oxidase | 4015 |
| 23 | HPRD_03123 | IL1RL1 | Interleukin 1 receptor-like 1 | 9173 |
| 24 | HPRD_09968 | G0S2 | G0/g1switch 2 | 50486 |
| 25 | HPRD_09545 | NREP | Neuronal regeneration related protein | 9315 |
| 26 | HPRD_00552 | NT5E | 5'-nucleotidase, ecto (CD73) | 4907 |
| 27 | HPRD_05081 | GPNMB | Glycoprotein (transmembrane) nmb | 10457 |
| 28 | HPRD_01903 | ITGAV | Integrin, alpha V | 3685 |
| 29 | HPRD_13735 | ILDR1 | Immunoglobulin-like domain containing receptor 1 | 286676 |
| 30 | HPRD_10420 | GFRA3 | GDNF family receptor alpha 3 | 2676 |
| 31 | HPRD_03115 | STC1 | Stanniocalcin 1 | 6781 |
| 32 | HPRD_04183 | IGFBP7 | Insulin-like growth factor binding protein 7 | 3490 |
| 33 | HPRD_16291 | PPP1R14A | Protein phosphatase 1, regulatory (inhibitor) subunit 14A | 94274 |
| 34 | HPRD_02698 | FABP4 | Fatty acid binding protein 4, adipocyte | 2167 |
| 35 | HPRD_01981 | PCCA | Propionyl coa carboxylase, alpha polypeptide | 5095 |
| 36 | HPRD_03774 | PER1 | Period circadian clock 1 | 5187 |
| 37 | HPRD_13516 | ZNF662 | Zinc finger protein 662 | 389114 |
| 38 | HPRD_06631 | MAGED4B | Melanoma antigen family D, 4B | 81557 |
| 39 | HPRD_11750 | GPX3 | Glutathione peroxidase 3 (plasma) | 2878 |

**Figure 1.**
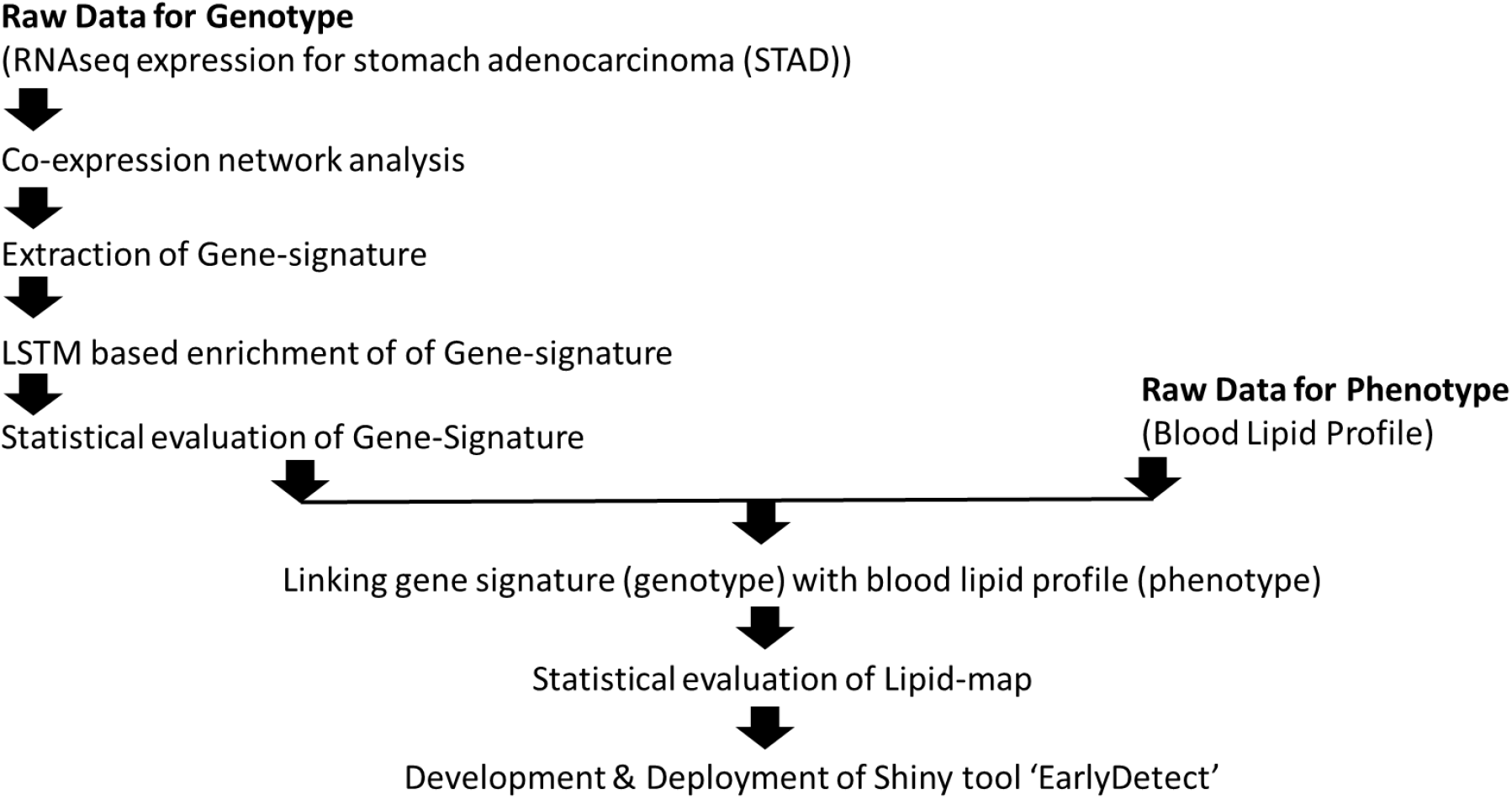
Workflow for Development of tool for early detection of stomach adenocarcinoma based on blood-lipid profile as clinicopathological feature

**Figure 2.**
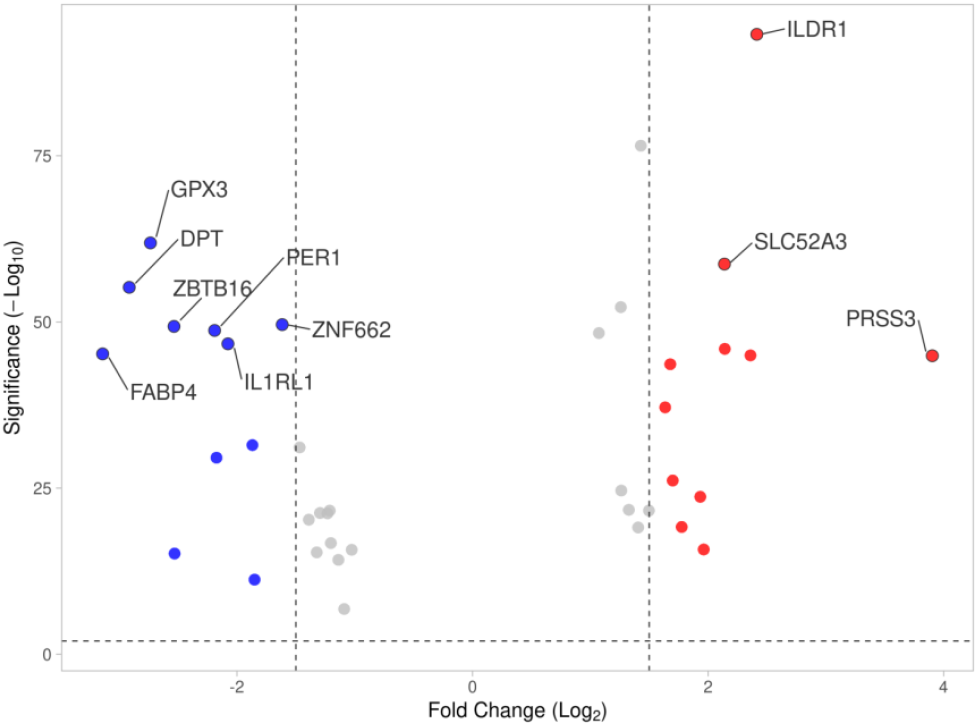
Volcano plot showing differentially expressed genes. Out of 4644 genes, 62 found to be differentially expressed. ‘Fold change (log_2_)’ threshold at 1.5. ‘Significance (-log_10_)’ boundary was 0.05.

### Gene-signature through network analysis

Gene co-expression network analysis performed to know the gene-signature for discrimination of population between disease-vs-normal. Considering 39 gene expression, signatures derived by using above methodology. Notation for Node-vs-Gene should be picked from table above. Two major results found: (A) Out of 39, only 11 genes found to be involved in defining signature through similarity matrix. Similarity network-based signature included AFF3, APBB1, C5, CHRD, COL4A5, EEF1A2, ZBTB16, IL1RL1, GFRA3, ZNF662, and MAGED4B genes; (B) Out of 39, only 10 genes found to be involved in defining signature through dis-similarity matrix. Dis-similarity network-based signature included AFF3, C5, CHAF1A, EFNA3, SLC52A3, LOX, ILDR1, GFRA3, STC1, and ZNF662. Four genes namely AFF3, C5, GFRA3 and ZNF662 were common in both the signatures. Both similarity & dissimilarity network signatures contained population discrimination capacity between normal & diseased (Figure 3). Survival & box plots showed gene-signatures identified (p-value < 0.05) with 11 genes extracted through network analysis (Figure 4). Survival plot validated the capacity of signature for significant discrimination between normal & diseased population.

**Figure 3.**
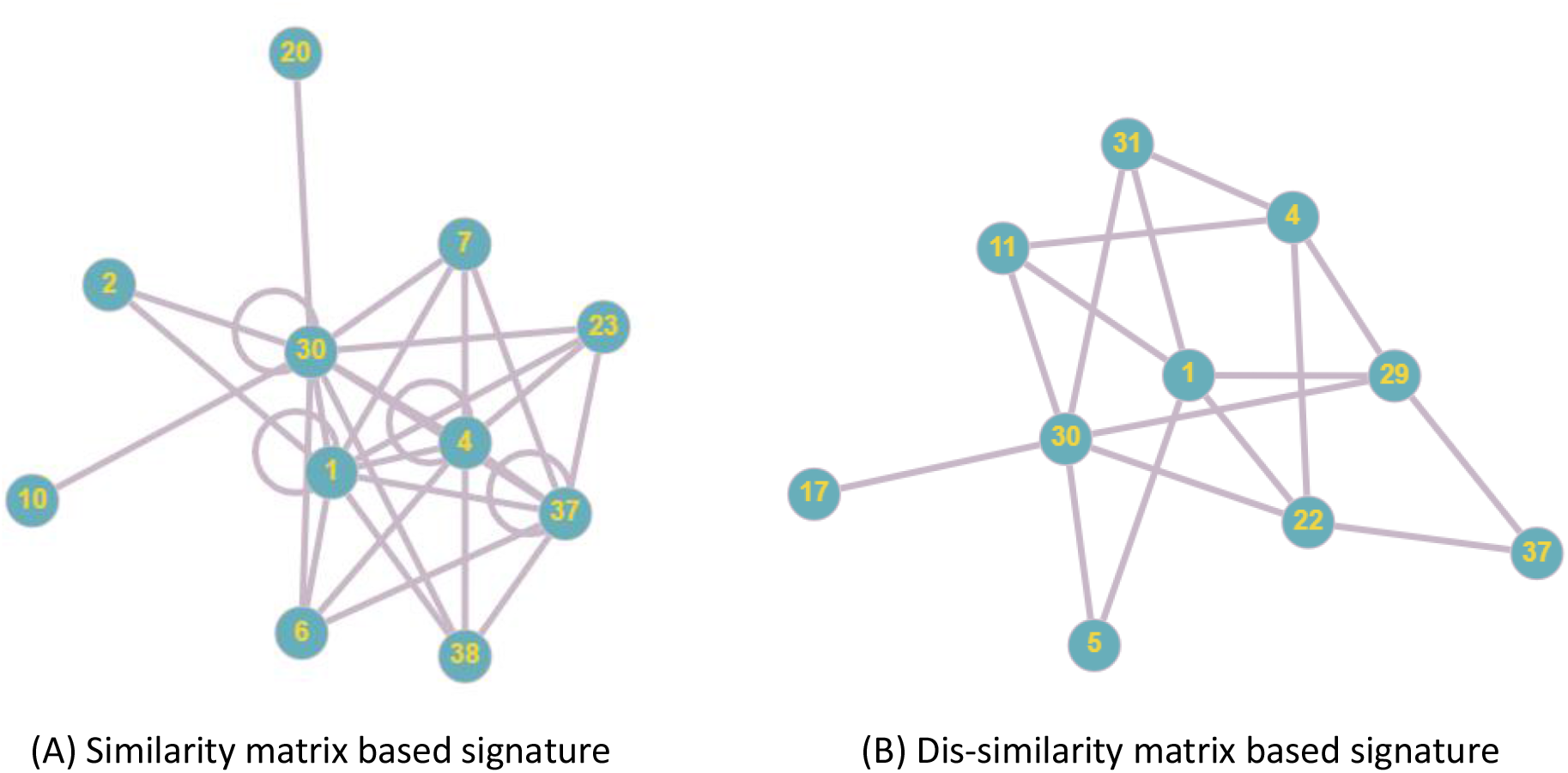
Considering 39 gene expression, signatures derived by the protocol described in above methodology. Signatures were extracted for discrimination between populations of diseased & non-diseased samples. Notation for Node-vs-Gene should be picked from table above. (A) Out of 39, only 11 genes were found to be involved in defining signature through similarity matrix. Similarity network-based signature included AFF3, APBB1, C5, CHRD, COL4A5, EEF1A2, ZBTB16, IL1RL1, GFRA3, ZNF662, and MAGED4B; (B) Out of 39, only 10 genes were found to be involved in defining signature through similarity matrix. Dis-similarity network-based signature included AFF3, C5, CHAF1A, EFNA3, SLC52A3, LOX, ILDR1, GFRA3, STC1, and ZNF662. Both signatures have common involvement of AFF3, C5, GFRA3 and ZNF662.

**Figure 4.**
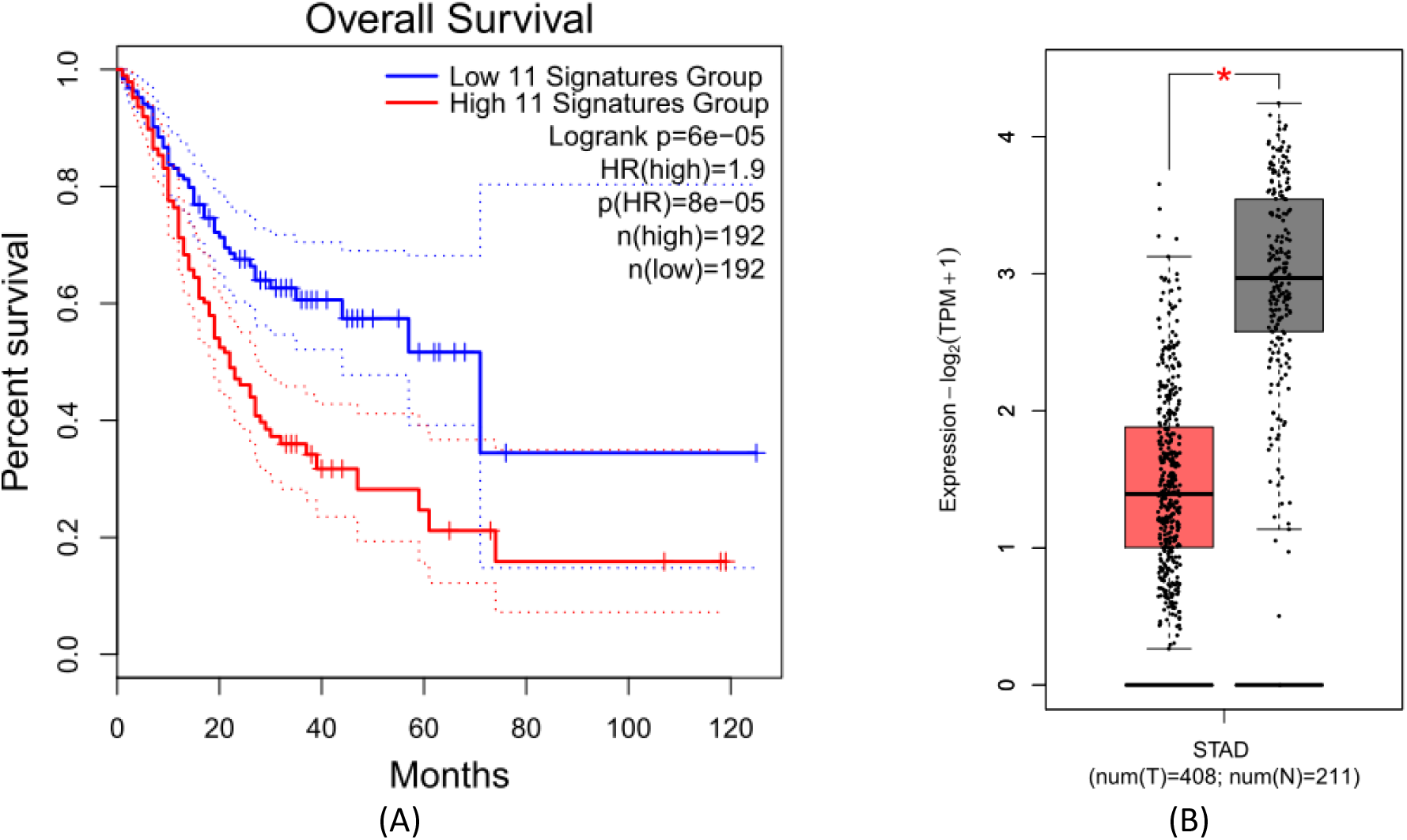
(A) Survival plots in reference of identified gene-signature (p-value < 0.05). Plot derived with 11 genes, extracted through network analysis; (B) Box plots showed the tissue-wise expression of multi-gene signature with 11 gene identified through network analysis.

### AI-guided Simulation of systems model & pathway enrichment

Similarity network (including 11 genes) became base for development of AI-guided systems models. Two system models developed; first for normal expression, and second for cancer (tumor) expression. AI-guided simulation of systems model resulted in only 05 genes approaching to stabilized state. Successful simulation involved 05 genes namely AFF3, APBB1, C5, CHRD, and COL4A5. Combination of simulated genes was available in both normal and tumor cases. Expression behavior on time scale also showed different patterns of expression between normal & diseased state (Figure 5(A&B)). Survival & box plots showed in reference of gene-signature identified (p-value < 0.05) with 05 genes extracted through enrichment through system model (Figure 6). It also validated significant discrimination between normal & diseased population. AFF3 gene is known to be a putative transcription activator, that may function in lymphoid development and oncogenesis. APBB1 gene is a Amyloid beta precursor, it role is in response to DNA damage & apoptosis. C5 gene is a Pyrimidine/ purine nucleoside phosphorylase. CHRD gene is a key development protien, and is used during early embryonic tissue and also expression cancer condition. COL4A5 gene is collagen alpha, it is used as constituent of extracellular matrix. Reactome based pathway enrichment analysis showed that, signatures genes belongs to the ‘Extracellular Matrix Organization’, ‘immune system’, ‘DNA repair’ and ‘developmental biology’. It is shown that most significant impact on stomach adenocarcinoma is due to genes involved in extracellular matrix organization. In a prior study, STAD early detection biomarker genes were suggested from extracellular matrix [30]. These observations are also in compliance of prior studies on development of gastric cancer [1] (Figure 7). These 05 gene were further used for establishing relation with blood profile.

**Figure 5.**
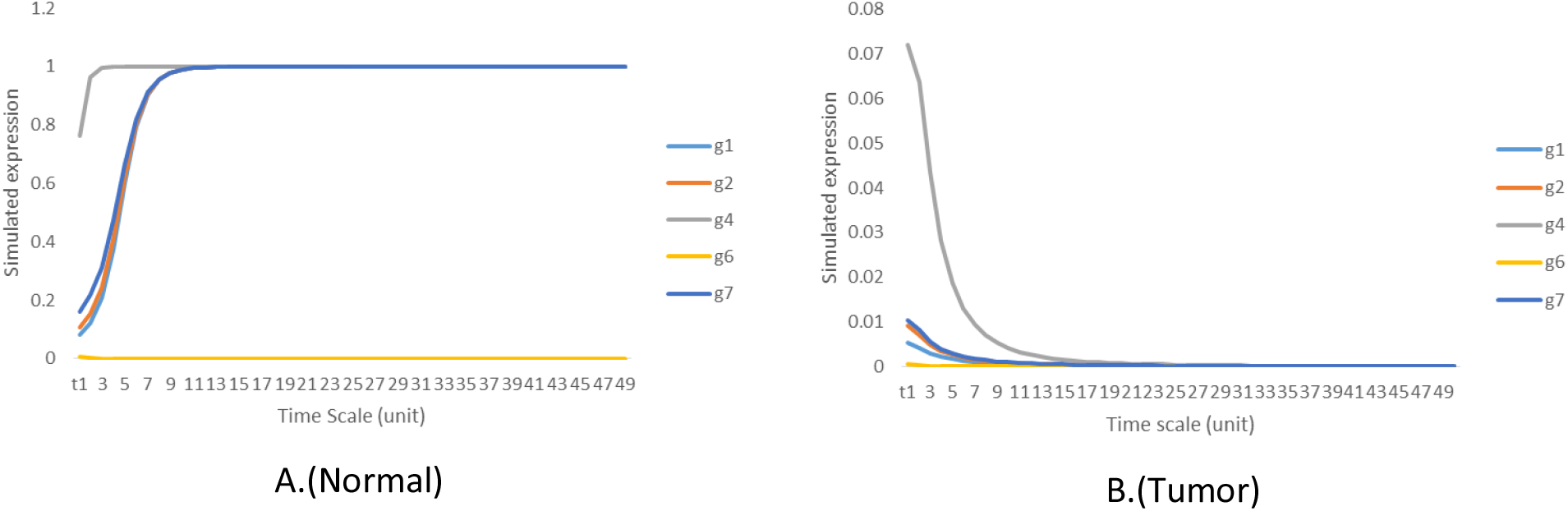
In present study, above similarity network-based signature (including 11 genes) used for development of AI-guided systems models. Two system models developed; first for normal expression, and second for cancer (tumor) expression. After simulation, it was found that out of 11 gene, only 05 genes (AFF3, APBB1, C5, CHRD, and COL4A5) were involved in successful simulation. Combination of these 05 gene were found in both normal and tumor cases. Above figures A&B showing the patterns of expression 05 gene; discrimination can be easily visualized between normal & tumor case. These expressions have been drawn on the time scale.

**Figure 6.**
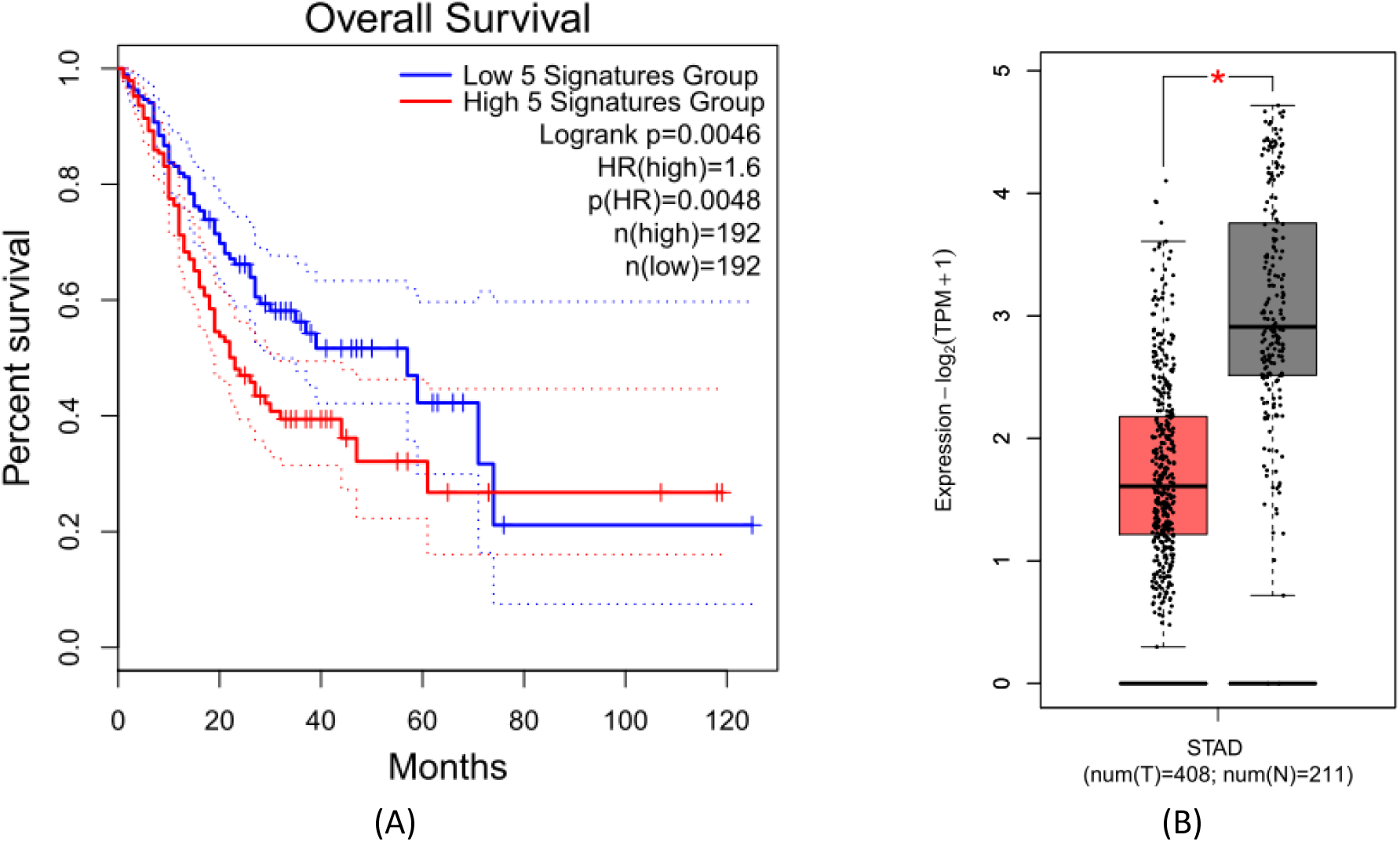
(A) Survival plot in reference of gene-signature identified (p-value = 0.0046 < 0.05). Plot derived with 05 genes extracted through system model based enrichment of similarity network; (B) Box plot showed the tissue-wise expression of multi-gene signature with 05 gene identified through enrichment of similarity network.

**Figure 7.**
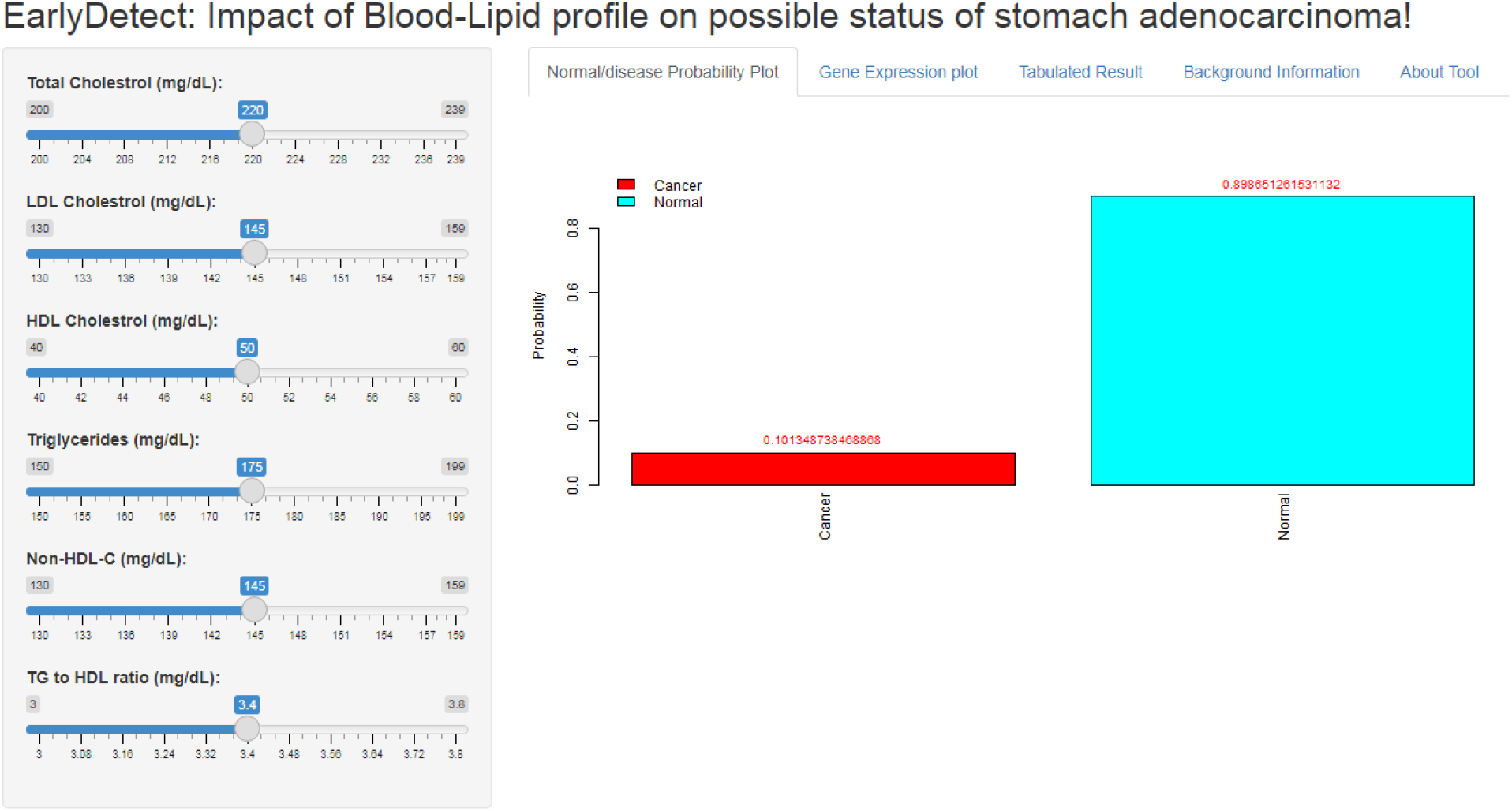
EarlyDetect tool description

### Linking gene signature (genotype) with blood lipid profile (phenotype)

Phenotype blood lipid profile used in this study contain: Total Cholestrol: 200-239(mg/dL); LDL Cholestrol: 130-159 (mg/dL); HDL Cholestrol: 40-60 (mg/dL); Triglycerides: 150-199 (mg/dL); Non-HDL-C: 130-159 (mg/dL); and TG to HDL ratio: 3.0-3.8 (mg/dL). Expression of gene signature further mapped with profile of blood. Normal-to-Cancer discrimination capacity of lipid profile was evaluated through multi-perceptron ANN classification model. Multiple perceptron model architecture of 6-4-2 implemented with learning rate 0.3 and momentum of 0.2 for 500 epochs. Model established with AUC of 0.999 (Table 2, 3, & 4). Furthermore, Genotype-to-Phenotype mapping resulted into ANN model. ANN model implemented in tool developed; where model performs reverse transformation of blood-profile into gene-expression.

**Table 2.**
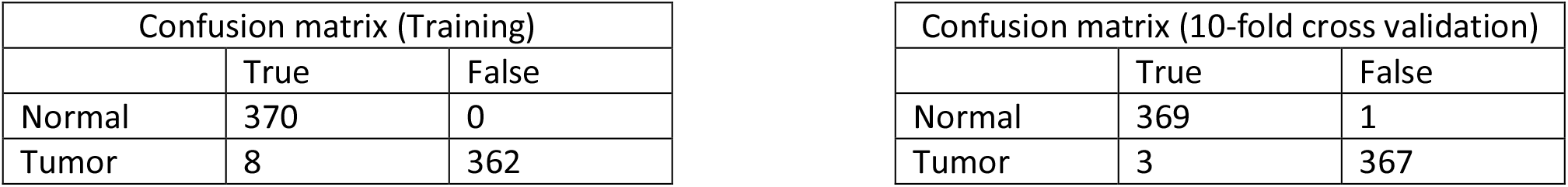
Confusion matrices showing discrimination capacity of lipid-profile performed by a two-class classification training model and 10-fold cross validation.

**Table 3.** Training statistics for two class-classification model for evaluation of discrimination capacity of lipid-profile.

|  | TP | FP | Precision | Recall | F-Measure | MCC | ROC area | PRC area | Class |
| --- | --- | --- | --- | --- | --- | --- | --- | --- | --- |
|  | 1 | 0.022 | 0.979 | 1 | 0.989 | 0.979 | 1 | 1 | T |
|  | 0.978 | 0 | 1 | 0.978 | 0.989 | 0.979 | 1 | 1 | N |
| Weighted Avg. | 0.989 | 0.011 | 0.989 | 0.989 | 0.989 | 0.979 | 1 | 1 |  |
| Correctly Classified Instances |  |  | 732 | 98.9189 % |  |  |  |  |  |
| Incorrectly Classified Instances |  |  | 8 | 1.0811 % |  |  |  |  |  |
| Kappa statistic |  |  | 0.9784 |  |  |  |  |  |  |
| Mean absolute error |  |  | 0.0116 |  |  |  |  |  |  |
| Root mean squared error |  |  | 0.0933 |  |  |  |  |  |  |
| Relative absolute error |  |  | 2.323 % |  |  |  |  |  |  |
| Root relative squared error |  |  | 18.6595 % |  |  |  |  |  |  |
| Total Number of Instances |  |  | 740 |  |  |  |  |  |  |

**Table 4.** Ten-fold cross-validation statistics for evaluation of two-class classification model showing discrimination capacity of lipid-profile.

|  | TP | FP | Precision | Recall | F-Measure | MCC | ROC area | PRC area | Class |
| --- | --- | --- | --- | --- | --- | --- | --- | --- | --- |
|  | 0.997 | 0.008 | 0.992 | 0.997 | 0.995 | 0.989 | 1 | 1 | T |
|  | 0.992 | 0.003 | 0.997 | 0.992 | 0.995 | 0.989 | 1 | 1 | N |
| Weighted Avg. | 0.995 | 0.005 | 0.995 | 0.995 | 0.995 | 0.989 | 1 | 1 |  |
| Correctly Classified Instances | 736 |  | 99.4595 % |  |  |  |  |  |  |
| Incorrectly Classified Instances | 4 |  | 0.5405 % |  |  |  |  |  |  |
| Kappa statistic | 0.9892 |  |  |  |  |  |  |  |  |
| Mean absolute error | 0.0091 |  |  |  |  |  |  |  |  |
| Root mean squared error | 0.0631 |  |  |  |  |  |  |  |  |
| Relative absolute error | 1.8151 % |  |  |  |  |  |  |  |  |
| Root relative squared error | 12.6272 % |  |  |  |  |  |  |  |  |
| Total Number of Instances | 740 |  |  |  |  |  |  |  |  |

### Development of Shiny tool ‘EarlyDetect’

The whole study resulted into ‘EarlyDetect’, which is a R-Shiny web-application. The tool is able to receive any combination of 06 parameters, as inputs of blood lipid profile, and can provide output in the form of probability of status of normal & cancer. It also provides information about the respective variation in gene-expression values in reference of lipid profile. Results can also be seen in tabulated format, including all the parameters & gene expressions. ‘EarlyDetect’ web application, process blood lipid profile through an ANN model to transform into gene-expression of signature. Further, featured gene expressions is used to calculate classified probability of class weightage of cancer-vs-normal. EarlyDetect predicts probability for both the classes (Figure 7).

### Utilization of ‘EarlyDetect’ for observation of parameter combinations on status of disease

The ‘EarlyDetect’ proved itself to be highly useful to observe the impact of various combinations of blood lipid profile in probability of cancer. Any combination of lipid profile parameters can be used, from patient, for analysis. Some examples are performed here as: (i) ***which parameter, at its lowest values, has maximum impact on inducing cancer***? To observe this situation, each parameter (one-by-one) was set up at lowest value, taking other parameters to default. It was observed that LDL has maximum impact (about 18%) on creating cancer condition. While probability of cancer further increases with increase in ‘TG to HDL ration’ parameter up to 20% (Figure 8). (ii) ***What may be the impact of total cholesterol on probability of cancer***? To observe this situation ‘total cholesterol value was started from lowest value of 200 mg/dL and increased step by step to maximum value of 239 mg/dL. At each step, probability of cancer was calculated. It was observed that probability of cancer decreases with increases of total cholesterol concentration. This observation was validated through a previous study [38] (Figure 9). It was found that with increase of ‘total cholestrol’, expression of AFF3, APBB1, C5 and CHRD increased, while expression of COL4A5 get decreased. (iii) ***What is biological relevance of decrease in expression of COL4A5 during increase of total cholestrol***? Decrease in expression of COL4A5 showed reduction in collagen. This observation was validated through a prior study [39] (Figure 10).

**Figure 8.**
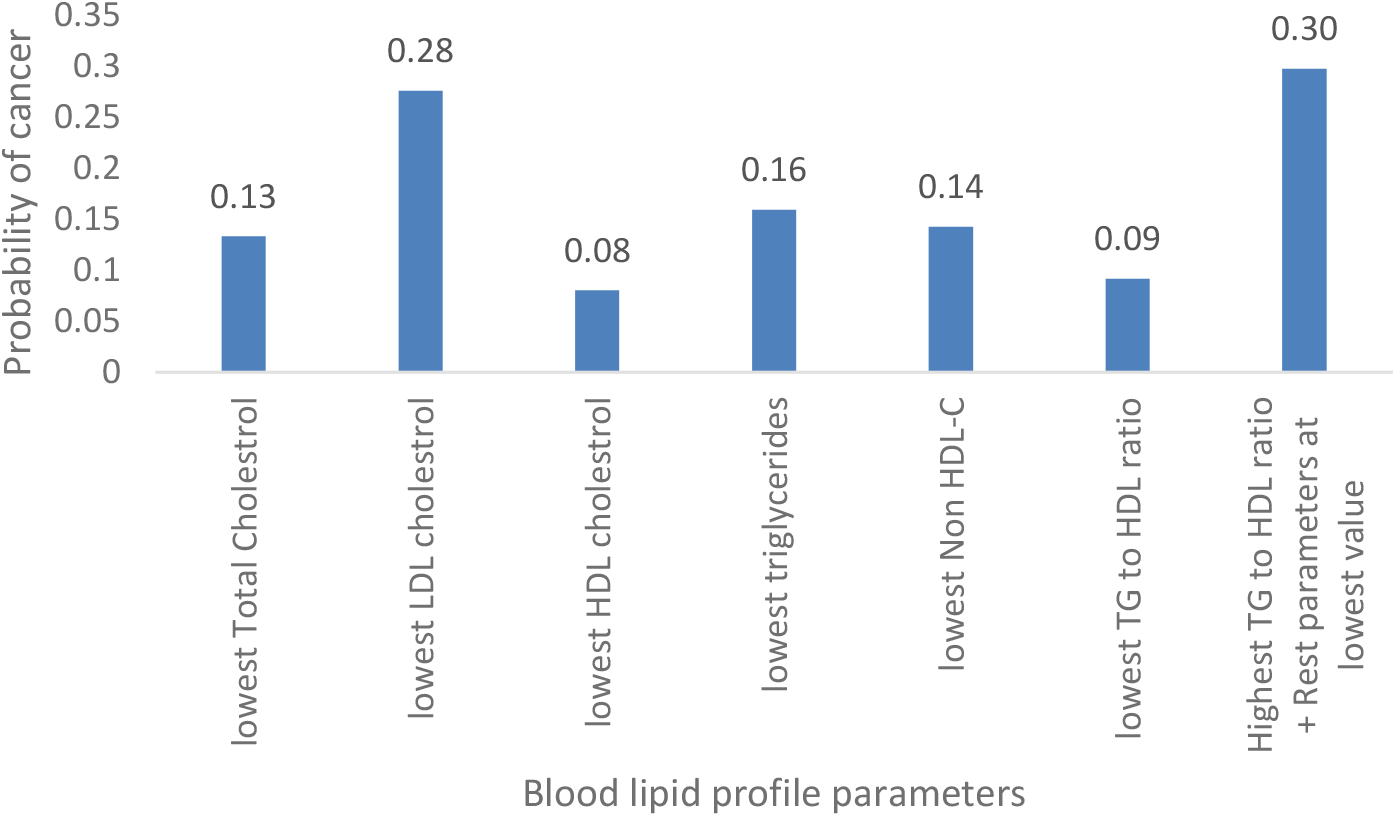
Behavior at the lowest value of each parameter

**Figure 9.**
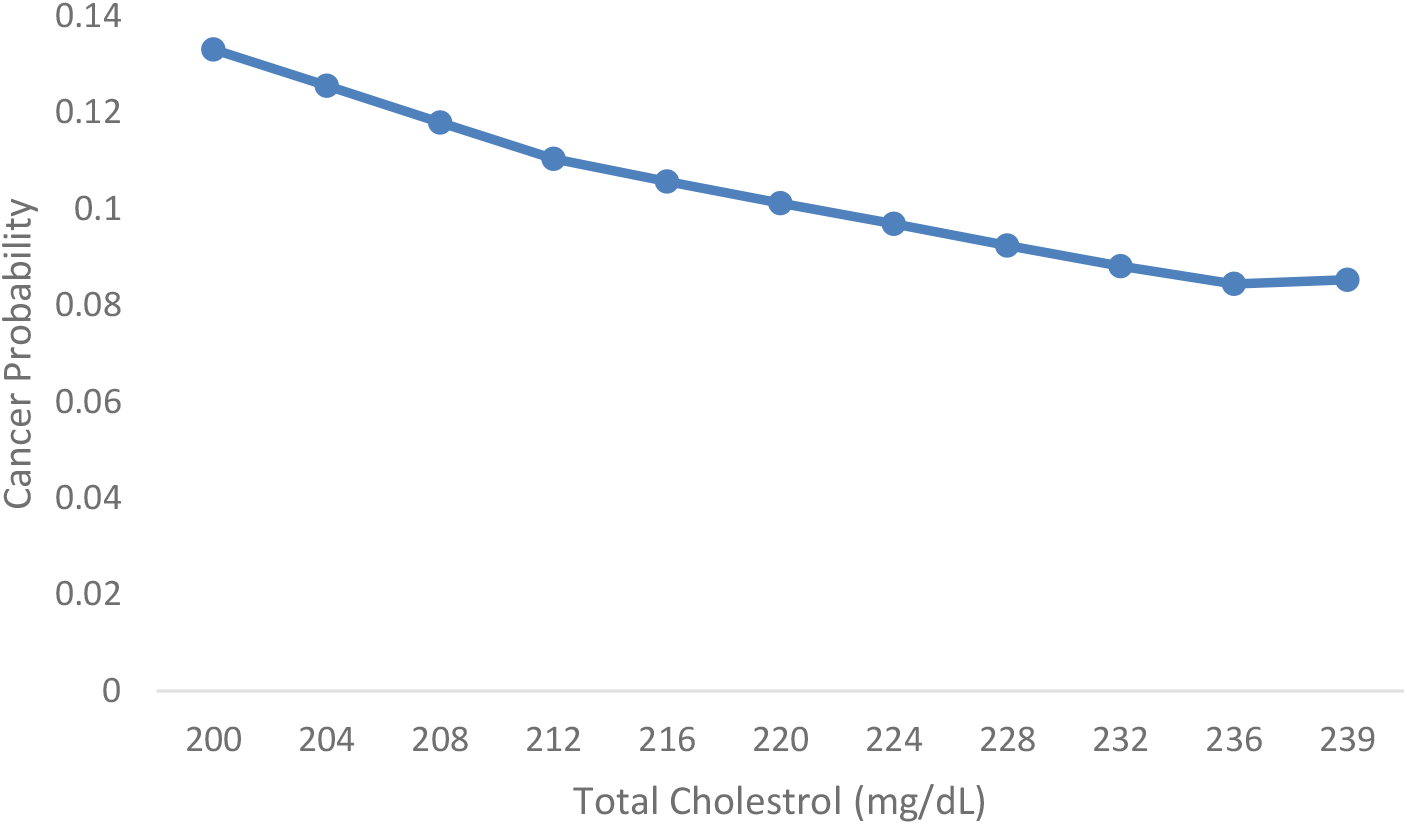
Cancer probability with increase of total cholesterol (mg/dL). Considering result of lipid parameters at their normal state

**Figure 10.**
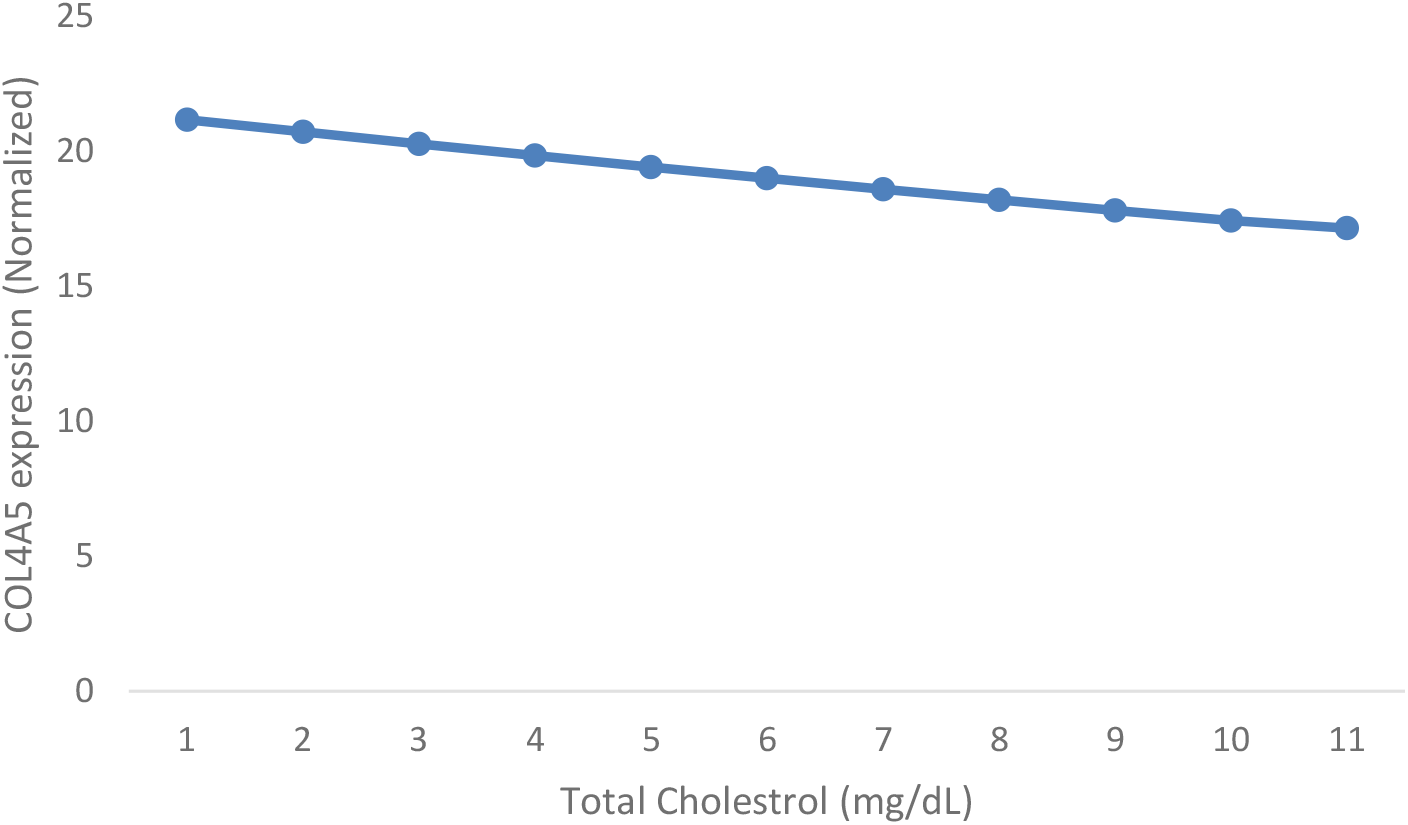
Expression of COL4A5 decreases with increase of total cholesterol (mg/dL). Considering result of lipid parameters at their normal state

## Conclusions

Present study successfully established the relationship between phenotype & genotype, as ‘blood profile’ & ‘gene signature’ respectively. This relationship was utilized for development of a tool for early detection of stomach adenocarcinoma on the basis of a clinicopathological feature blood-lipid profile. Few identified conclusions based on EarlyDetect tool are as: (i) Low-Density-Lipoprotein cholesterol showed potential impact on creating cancer; (ii) Probability of cancer decreases with increase in total cholesterol; (iii) Collagen decreases with increase in total cholesterol. More conclusions can be drawn through combination studies through ‘EarlyDetect’. Tool is freely accessible at ‘https://csir-icmr.shinyapps.io/EarlyDetect/’.

## Data Availability

All data produced in the present work are contained in the manuscript.

## Availability and requirements

**Project name**: ICMR-RA (IRIS no.: 2020-5987)

**Project home page**: https://csir-icmr.shinyapps.io/EarlyDetect/

**Operating system(s)**: Platform independent (Online web-Browser).

**Programming language**: R

**Other requirements**: Internet access.

**License**: GNU.

**Any restrictions to use by non-academics**:

No restriction.

## Acknowledgement

Authors are thankful to the Director, CSIR-Central Institute of Medicinal & Aromatic Plants (CIMAP), Lucknow, India for infrastructure & research facilities support. Author OP is thankful to the Indian Council of Medical Research (ICMR), New Delhi, India for financial support through RA fellowship (Award letter no. BMI/11(12)/2020, dated: 04/02/2021). Author OP is also thankful to Prof. Thiol Gross, Helmholtz Institute for Functional Marine Biodiversity (HIFMB) at University of OLDENBURG, Germany and Dr. Amit Singh, Chennai Mathematical Institute (CMI) Chennai, India for fruitful suggestions on evaluation of stability of time series systems model. The CSIR-CIMAP publication number of this manuscript is CIMAP/PUB/ 2022/65.

